# Next-generation phenotyping in Nigerian children with Cornelia de Lange Syndrome

**DOI:** 10.1101/2024.02.15.24302695

**Authors:** Annabelle Arlt, Alexej Knaus, Tzung-Chien Hsieh, Hannah Klinkhammer, Meghna Bhasin, Alexander Hustinx, Shahida Moosa, Peter Krawitz, Ekanem Ekure

## Abstract

Next-generation phenotyping (NGP) can be used to compute the similarity of dysmorphic patients to known syndromic diseases. So far, the technology has been evaluated in variant prioritization and classification, providing evidence for pathogenicity if the phenotype matched with other patients with a confirmed molecular diagnosis.

In a Nigerian cohort of individuals with facial dysmorphism, we used the NGP tool GestaltMatcher to screen portraits prior to genetic testing and subjected individuals with high similarity scores to exome sequencing (ES). Here, we report on two individuals with global developmental delay, pulmonary artery stenosis, and genital and limb malformations for whom GestaltMatcher yielded Cornelia de Lange syndrome as the top hit. ES revealed a known pathogenic nonsense variant, NM_133433.4: c.598C>T; p.(Gln200*), as well as a novel frameshift variant c.7948dup; p.(Ile2650Asnfs*11) in *NIPBL*. Our results suggest that NGP can be used as a screening tool and thresholds could be defined for achieving high diagnostic yields in ES. Training the AI with additional cases of the same ethnicity might further increase the positive predictive value of GestaltMatcher.

## Introduction

GestaltMatcher is an artificial intelligence (AI) that has been developed to analyze phenotypic features in medical imaging data of patients with rare diseases (Hsieh et al., 2022). The technology has been evaluated in a prospective setting in a large study of the national healthcare system in Germany and shown to increase the diagnostic yield of exome sequencing (Schmidt et al., 2023). In that study, a multidisciplinary team (MDT) of clinicians had to assess the clinical features of patients who were suspected of having rare disorders. If the MDT concluded that a genetic cause was likely, the patients were subjected to exome sequencing (ES), and the results of the GestaltMatcher analysis were used in the data analysis (Hsieh et al., 2019). We hypothesized that gestalt scores could be used not only in variant prioritization and classification but also in the decision-making process prior to diagnostic testing. We, therefore, computed the positive likelihood ratio (LR^+^) for different disorders based on the GestaltMatcher scores and defined thresholds suggesting a high positive predictive value (PPV) (Lesmann et al., 2023). We identified two Nigerian patients submitted to GestaltMatcher who scored high for Cornelia de Lange syndrome and for whom no genetic testing had yet been performed.

Cornelia de Lange syndrome (CdLS, OMIM #122470, 300590, 610759, 300882, and 614701) is an autosomal dominant multisystem malformation syndrome with an estimated incidence of 1:10,000 to 1:30,000 live births (Mannini et al. 2013). CdLS can be caused by pathogenic variants in the regulatory subunits of the cohesin complex encoded by *NIPBL* and *HDAC8* or in the structural subunit of *SMC1A, SMC3*, and *RAD21*.

*NIPBL* is the major CdLS disease gene, and heterozygous variants are identified in more than 65% of affected individuals (Musio et al. 2006; Rohatgi et al. 2010). CdLS is recognized by characteristic facial dysmorphism, including low anterior hairline, arched eyebrows, synophrys, anteverted nares, maxillary prognathism, long philtrum, thin lips, and ‘carp’ mouth, in association with prenatal and postnatal growth retardation, developmental delay and upper limb anomalies.

Few publications have focused on syndromes in diverse populations (Dowsett et al. 2019). As a result, many clinicians are trained with clinical genetic resources where only individuals of European descent serve as the standard of reference (Muenke, Adeyemo, and Kruszka 2016). Here, we compare facial dysmorphism and findings from physical exams in individuals from underrepresented minority groups with CdLS and demonstrate how facial analysis technology can be a useful clinical tool in the diagnosis of individuals from diverse ancestral backgrounds and developing countries. Next-generation phenotyping approaches (NGP), such as GestaltMatcher (Hsieh et al. 2022), have been proven capable of facilitating the diagnosis of rare disorders (Forwood et al. 2023; Schmidt et al. 2023). However, they have not yet been evaluated systematically on ethnically diverse datasets, as many populations are still disproportionately underrepresented, notably Africans. In this study, we show that GestaltMatcher can predict *NIPBL*, the disease-causing gene, correctly on the first rank for both individuals. The clinical diagnosis was molecularly confirmed by trio exome sequencing analysis. We demonstrated the effectiveness of the NGP approach on two Nigerian patients. Moreover, utilizing this technology is crucial to facilitate the diagnostic process and to reduce the cost to the healthcare system in developing countries with limited or no access to specialist medical geneticists.

## Patients, materials and methods Clinical details

### Patient 1 (GMDB Case ID: 9577)

A boy with global developmental delay, microcephaly, growth delay, cryptorchidism, micropenis and pulmonary artery stenosis was seen for syndromal diagnostics. Craniofacial dysmorphic features included synophrys, long eyelashes, thick hair, depressed nasal bridge, and low anterior hairline. He also presents bilateral postaxial hand polydactyly. He is the second child born to non -consanguinous healthy parents. The older sibling is male and has no obvious abnormality. Pregnancy history was unremarkable. He has had balloon valvuloplasty for the pulmonary stenosis and ochiopexy for cryptorchidism.

### Patient 2

The second boy presented with global developmental delay, intellectual disability, growth delay and cryptorchidism. He had dysmorphic facial features typical for CdLS, including synophrys, highly arched eyebrow, long eyelashes, facial hirsutism, hypertelorism, epicanthus, depressed nasal bridge, long and smooth philtrum, low anterior hairline, low-set ears, anteverted nares, thin upper and lower lip vermilion, prominence of the premaxilla and a high palate. He also had split hand deformity. Moreover, no visual contact was possible. He was born to non-consanguinous healthy parents after 38 weeks gestation. Pregnancy was uneventful but baby was smaal for gestational age with birth weight of 1.8 kg. Family history was unremarkable.

## Materials and Methods

DNA was extracted from saliva and/or blood samples, enriched with TWIST Exome CoreRefSeq, and sequenced with a 100 bp paired-end protocol on a NovaSeq 6000. Sequence reads were mapped to the reference genome GRCh37 with BWA-MEM, and variants were called with GATK according to best practice guidelines. 95% of the target region was covered by at least 20x. We utilized GestaltMatcher to analyze the patient’s facial image (Hustinx et al. 2022, Hsieh et al., 2023). The facial image is first encoded by the GestaltMatcher model into the facial phenotype descriptor. We then calculated the cosine distance between the test image to 9,764 facial images of patients with 528 different rare disorders in the GestaltMatcher Database (GMDB) (Lesmann et al. 2023). The suggested disease-causing gene list was further ranked by sorting the cosine distances in the accessing order. Variant analysis was done using the prioritization of exome data by image analysis (PEDIA) approach (Hsieh et al. 2019) in the VarFish platform (Holtgrewe et al. 2020). First, phenotypic similarities of clinical features to rare disorders were assessed by CADA with the Human Phenotype Ontology (HPO) terminology (Peng et al. 2021). We then quantified the facial gestalt scores by analyzing the facial image with GestaltMatcher. On the molecular level, the deleteriousness of pre-filtered variants was assessed by CADD scores (Kircher et al. 2014), and the highest score per gene was annotated. All scores were used to compute a PEDIA score per gene, and genes were ranked by the PEDIA score in descending order. The final output of the PEDIA score for each variant can be utilized for prioritization. Further evaluation was carried out using VarFish11. The identified sequence variants were assessed on the basis of public databases (Deciper (https://decipher.sanger.ac.uk/), ClinVar (https://www.ncbi.nlm.nih.gov/clinvar/), 1kGP12, Iranome13, GME Variome14 and gnomAD (http://gnomad.broadinstitute.org/). Recommendations for the classification of variants according to ACMG guidelines were also taken into account. Non-pathogenic or low-penetrating polymorphisms (pathogenicity classes 1 and 2 according to ACMG/AMP guidelines) are not documented in this finding. Variants of unclear significance (pathogenicity classes 3 and 4 according to ACMG/AMP guidelines) are only mentioned if, based on the literature, a (partial) causation of the variant for the individual’s clinical situation is considered possible. If necessary, a full list of the identified variants can be requested. The assessment of the clinical relevance of identified variants may change in the future due to scientific progress. Variants that are not compatible with a plausible inheritance (autosomal recessive or autosomal dominant) assuming complete penetrance were not considered. Any additional findings can only be made in the genes analyzed; additional findings include variants in disease genes that are relevant for preventive care or therapy. Carriers, e.g. for autosomal recessive diseases, are only reported if a (partial) causation of the variant to the individual’s clinic is considered possible. Variants with a population frequency of 3% in population databases were not considered. Intronic variants (beyond ± 20bp of the splice point) and variants in regulatory areas were not examined.

## Results

### Facial analysis technology and molecular genetic findings

#### Patient 1

We first analyzed a portrait image of individual 1 with GestaltMatcher, and CdLS was ranked in the top-1 position, indicating the most likely diagnosis based on syndromic similarity. In fact, the ten most similar patients in GMDB had a pathogenic variant in *NIPBL*, amongst them five of African ethnicity. Moreover, Gradient-weighted Class Activation Mapping (Grad-CAM) (Selvaraju et al. 2017) also indicated a depressed nasal bridge and typical synophrys, which are relevant facial dysmorphic features to CdLS.

By trio exome sequencing, we identified a novel pathogenic *de novo* frameshift variant in *NIPBL*, NM_133433.3:c.7948dup (p.Ile2650Asnfs*11), establishing the diagnosis of CdLS. The heterozygous duplication was covered with 230 reads, shows an alternate allele balance of 0.36 (ref: 229 reads C; alt: ins A 83 reads), and was only observed in the index while the parents are homozygous for the reference allele. This variant is therefore considered a *bona fide de novo* variant. The single base pair duplication c.7948dup leads to an exchange of the highly conserved amino acid isoleucine to asparagine at position 2650 following a frameshift and a premature stop codon after 11 amino acids in *NIPBL*. Bioinformatic prediction by MutationTaster and UMD Predictor classified this variant as pathogenic. Using PEDIA, the disease-causing variant in *NIPBL* was prioritized at rank 1.

On the basis of the clinical findings, facial analysis, *de novo* mutation status, and impact on RNA and protein level (loss of function mutation), we classify the novel variant c.7948dup p.(Ile2650Asnfs*11) in *NIPBL* as disease-causing in the affected individual.

#### Patient 2

For individual 2, GestaltMatcher also ranked *NIPBL* in the top-1 position, which was based on the first nine most similar patients in GMDB with this molecular diagnosis. Six of these patients were also of African ethnicity. Trio exome sequencing confirmed CdLS1 for case 2 and revealed a heterozygous *de novo* nonsense variant NM_133433.4:c.598C>T (p.Gln200*) in *NIPBL*. The heterozygous variant was covered with 89 reads, shows an alternate allele balance of 47% (ref: 47 reads C; alt: T 42 reads), was only observed in the index, while the parents are homozygous for the reference allele, and therefore is considered a bona fide *de novo* variant. The single base exchange c.598C>T leads to an exchange of a highly conserved amino acid glutamine to a stop codon at position 200 in *NIPBL*. Bioinformatic prediction by MutationTaster and UMD Predictor classified this variant as pathogenic with a CADD score of 37. The variant is listed as pathogenic in ClinVar: Accession ID: VCV000159167.2; Variation ID: 159167. Using PEDIA, *NIPBL* was prioritized on position 1.

On the basis of the clinical findings, facial analysis, *de novo* mutation status, and impact on RNA and protein level (loss of function mutation), we also classify the variant c.598C>T p.(Gln200*) in *NIPBL* as disease-causing in the affected individual.

## Discussion

CdLS is a syndromic disorder with a highly distinctive facial gestalt that can be learned and recognized by an AI, such as GestaltMatcher (Hsieh et al., 2019). Depending on the measured similarity the gestalt score can provide supporting or even moderate evidence for the pathogenicity of variants that are identified in the genes associated with this phenotype (Lesmann et al., 2023; Tavtigian et al., 2018). Similarly to the Bayesian reasoning in variant classification, a threshold for the similarity to CdLS could be used prior to genetic testing to adjust the expected diagnostic yield, especially in situations where resources are limited or costs need to be strictly managed. We tested this approach in a cohort of cases with facial dysmorphism that was collected in Nigeria. We identified two individuals with gestalt scores of 0.58 and 0.49 for CdLS and submitted samples for genetic testing. Trio exome analyses established the molecular diagnoses by revealing pathogenic *de novo* variants in *NIPBL*. It is noteworthy that the strong phenotypic match could only be achieved due to molecularly solved cases that were already available in the GestaltMatcher training set. Currently, only 4% of all individuals in the GestaltMatcher database are of African descent. If only CdLS patients of European ethnicity had been used for training the AI, the measured phenotypic similarity and therefore the achieved evidence of pathogenicity would have been lower (Figure 1). This underlines the importance of collecting and sharing medical imaging data of patients from ethnicities currently underrepresented in data collections for AI.

**FIGURE 1:**
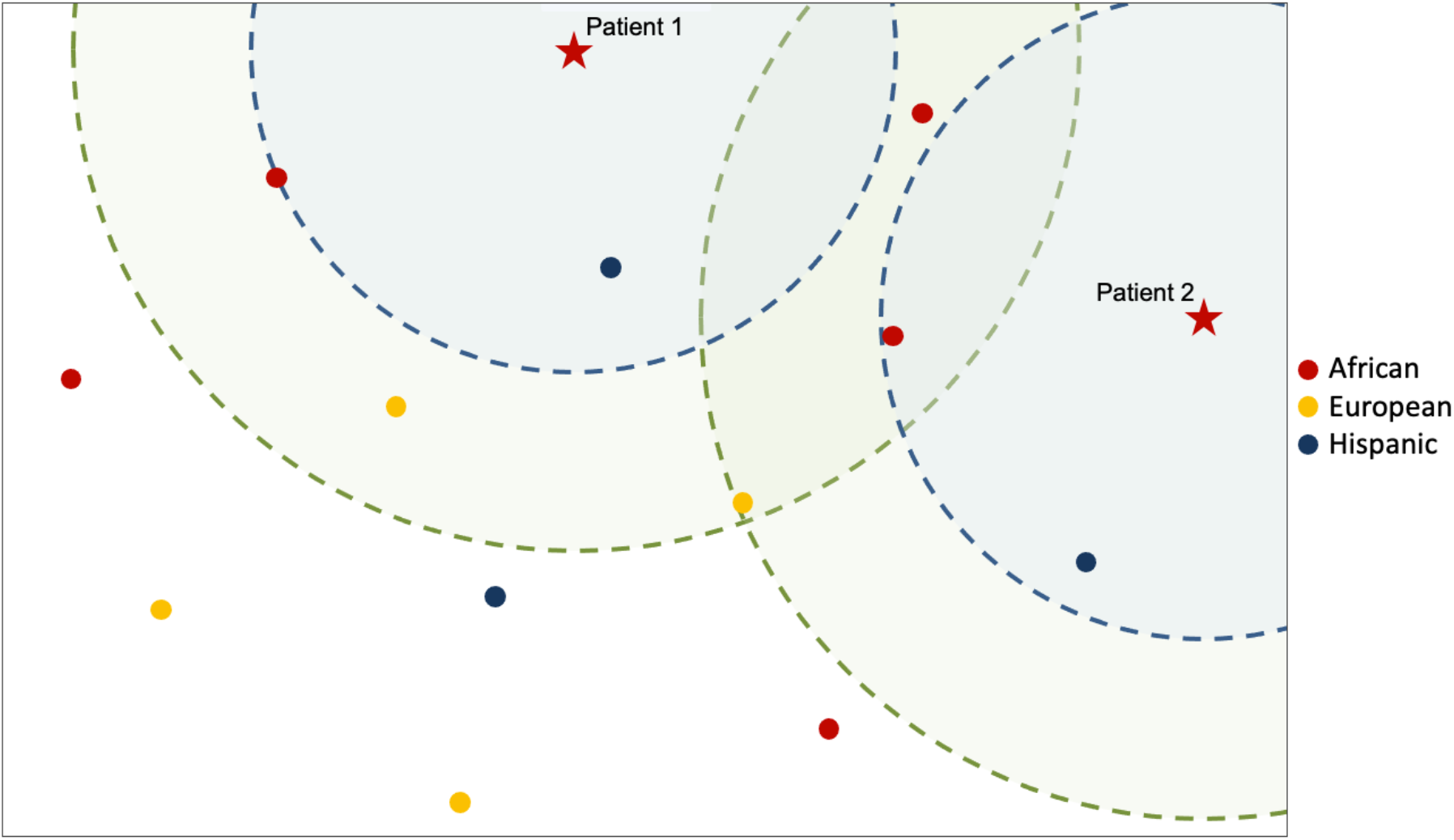
Two-dimensional representation of the facial phenotype descriptors of 13 CdL patients computed via GestaltMatcher, including the two presented patients (marked as stars) and their ten closest CdL matches, respectively. Blue circles indicate that CdL patients of African or Hispanic ancestry were closest to the two tested patients, whereas patients of European ancestry could only reach a weaker phenotypic similarity (indicated by green circles). In the Bayesian reasoning, this suggests that by including individuals of non-European ancestry, the positive predictive value of a phenotypic match can be substantially increased.

## Acknowledgments

We acknowledge the NGS Core Facility for their expertise in ES data generation and the CUBA for their proficient data processing, both of which were integral to this study’s success. Their contributions have been indispensable in advancing our scientific understanding. And the authors thank the family members who participated in this study.

## Ethics approval and consent to participate

Informed consent was obtained from all patients and the ethics committee of University of Lagos had granted ethical approval.

## Consent for publication

Legal guardians of patients gave written consent to use the photographs presented for publication.

## Data availability

The variants have been submitted to ClinVar (Accession: VCV001328515.1 and VCV000159167.6)

